# Prognostic impact of atrial function in cardiac sarcoidosis using two-dimensional speckle-tracking echocardiography

**DOI:** 10.1101/2023.12.15.23300054

**Authors:** Takahiro Nishihara, Yoichi Takaya, Rie Nakayama, Yu Yoshida, Norihisa Toh, Kazufumi Nakamura, Shinsuke Yuasa

**Affiliations:** Department of Cardiovascular Medicine, Okayama University Graduate School of Medicine, Dentistry and Pharmaceutical Sciences, Okayama, Japan

**Keywords:** atrial function, cardiac sarcoidosis, prognosis, echocardiography

## Abstract

**Aims:** Although ventricular dysfunction is associated with adverse outcomes in cardiac sarcoidosis (CS), the potential role of atrial function is unknown. The aim of this study was to assess the effect of atrial function on clinical outcomes in patients with CS.

**Methods:** We retrospectively enrolled 96 patients with CS. Left atrial (LA) and right atrial (RA) function was calculated as LA global longitudinal strain (LAGLS) and RA global longitudinal strain (RAGLS), respectively, using two-dimensional speckle-tracking echocardiography. Major adverse cardiac event (MACE) was a composite of cardiac death, fatal ventricular arrhythmia events, and hospitalization for heart failure.

**Results:** During a median follow-up of 6.0 years, 37 patients had MACE. LAGLS and RAGLS were lower in patients with MACE than in those without MACE. Kaplan-Meier curves showed that patients with atrial dysfunction with LAGLS of ≤ 19.6% or RAGLS of ≤ 22.3% had a higher rate of MACE than those without atrial dysfunction (log-rank test, p = 0.01 for both comparisons). The presence of LAGLS of ≤ 19.6% or RAGLS of ≤ 22.3% was significantly associated with MACE in a model that included age, sex, and New York Heart Association class (hazard ratio: 2.19, 95% confidence interval: 1.06–4.55, p = 0.04; hazard ratio: 2.27, 95% confidence interval: 1.07–4.85, p = 0.03, respectively).

**Conclusions:** Atrial dysfunction represented by LAGLS and RAGLS is associated with adverse outcomes in patients with CS. Our findings suggest a potential role of atrial function for predicting the prognosis in CS.

## Introduction

Sarcoidosis is a systemic inflammatory granulomatous disease of unknown etiology that can affect any organ. Approximately 25% of individuals who are diagnosed with systemic sarcoidosis have been reported to have cardiac involvement ^1^. Since cardiac involvement is a major contributor to mortality in patients with sarcoidosis, early recognition of cardiac sarcoidosis (CS) is crucial ^2–5^. CS impairs left ventricular (LV) contractility, leading to congestive heart failure and ventricular tachyarrhythmias. The extent of myocardial damage is an important predictor of adverse outcomes ^6^.

Sarcoidosis affects various lesions in the heart. Recent advances in imaging technologies have shown that right ventricular (RV) dysfunction and RV late gadolinium-enhancement are associated with adverse cardiac events ^7^ ^8^. Cardiac involvement of sarcoidosis occurs in the atrium as well as in the ventricle. Inflammation and scarring cause atrial remodeling, which progresses the impairment of atrial function ^9^. Atrial dysfunction may lead to worse clinical outcomes during long-term follow-up in patients with CS. However, despite evidence regarding ventricular function, the relationship between atrial function and the prognosis in patients with CS has not been assessed. In recent years, two-dimensional speckle-tracking echocardiography (2D-STE), which is a non-invasive and automatic method that tracks specified myocardial speckles to evaluate myocardial motion, has enabled the assessment of atrial function. Therefore, this study aimed to investigate the effect of atrial function using 2D-STE on clinical outcomes in patients with CS.

## Methods

### Study population and design

We retrospectively enrolled 100 patients who were diagnosed with CS at Okayama University Hospital, Japan from May 2008 to January 2023. We excluded patients with chronic atrial fibrillation because of insufficient tracking of the atrial walls by 2D-STE (n = 1) and less than one year of follow-up (n = 3). Figure 1 shows a flow diagram of the study design. Finally, 96 patients were included in this study. These patients were diagnosed by the Japanese Ministry of Health and Welfare criteria modified in 2006 ^10^ and the Japanese Circulation Society criteria proposed in 2016 ^11^. Baseline characteristics, such as age, sex, comorbidities, drug therapy, blood test data, and transthoracic echocardiographic findings, were obtained from medical records. Baseline was defined as the time that a patient underwent transthoracic echocardiography at our hospital for the first time.

**Figure 1.**
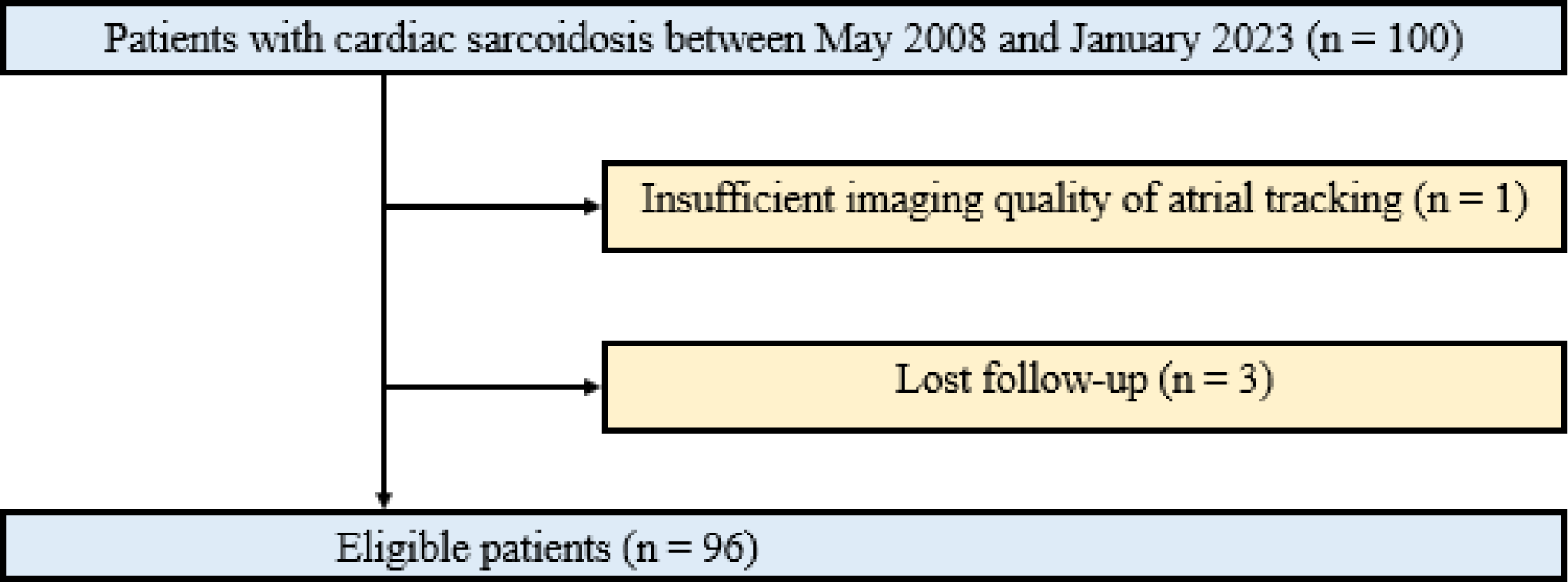
The study flow diagram.

This study was conducted by following the principles of the Declaration of Helsinki. The ethics committee of the Okayama University Graduate School of Medicine, Dentistry, and Pharmaceutical Sciences approved the study. Informed consent was waived because of the retrospective and low-risk nature of the study.

### Echocardiographic examination

Transthoracic echocardiography was performed using commercially available equipment (iE33; Philips Medical Systems, Andover, MA, USA, Vivid E9 and E95; GE Healthcare, Milwaukee, WI, USA, and Aplio i900; Canon Medical Systems, Otawara, Japan). Echocardiographic measurements were performed according to the guidelines ^12^ ^13^. LV volume and LV ejection fraction were calculated using the disc summation technique. Early and late peak diastolic velocities were measured using pulsed-wave Doppler. Tissue Doppler-derived early diastolic mitral annular velocity was measured at the septal and lateral wall sites. The ratio of early diastolic transmitral inflow velocity to early diastolic mitral annular velocity was calculated.

### Speckle-tracking strain analysis

Atrial function of the left atrium (LA) and the right atrium (RA) was analyzed using vendor-independent software (TOMTEC Imaging System, Unterschleissheim, Germany) (Figure 2). All echocardiographic images were sent in DICOM format to the core laboratory. Atrial global longitudinal strain (GLS) was quantified using apical four-chamber view images. Three reference points were placed at the mitral and tricuspid annulus and the roof of the atrium. The software automatically determined the atrial endocardial border and performed speckle-tracking analysis through one complete cardiac cycle. When an inaccurate atrial endocardial border was detected, manual adjustment was performed to ensure optimal tracking. Two dimensional-STE provided values of the average GLS of the LA and RA (LAGLS and RAGLS, respectively), LA and RA volume index, LA and RA fractional area change, and LA and RA ejection fraction. RV fractional area change was also measured from RV-focused apical four-chamber view. One experienced cardiologist who was blinded to clinical information performed the analyses of 2D-STE.

**Figure 2.**
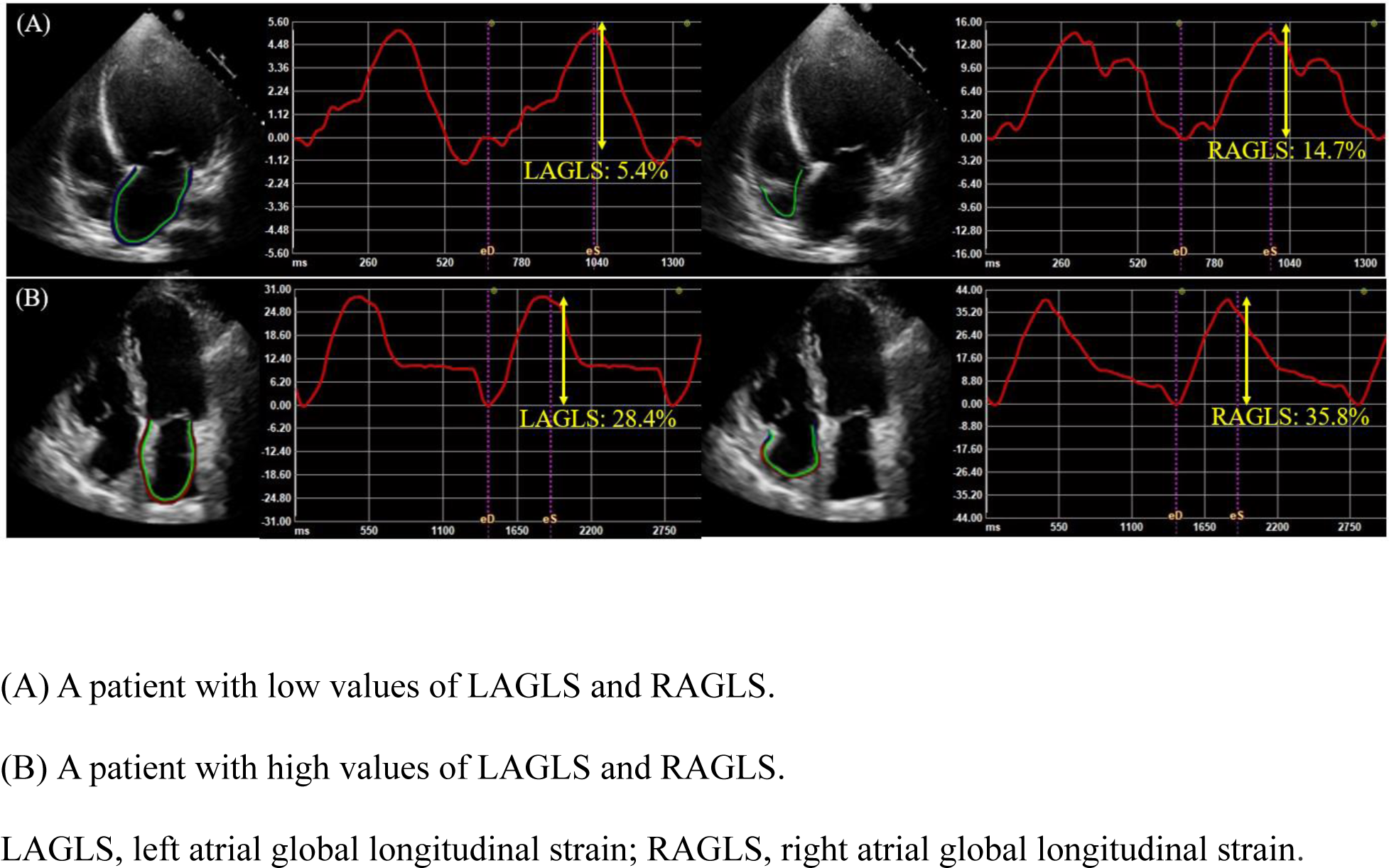
Representative cases of LAGLS and RAGLS.

### Clinical outcomes

Major adverse cardiac event (MACE) was a composite outcome of cardiac death, fatal ventricular arrhythmia events, and hospitalization for heart failure. Fatal ventricular arrhythmia events were defined as a combination of documented ventricular fibrillation and sustained ventricular tachycardia lasting > 30 s, or appropriate implantable cardioverter defibrillator therapy. All outcome data were retrospectively obtained from medical records and telephone interviews.

### Statistical analysis

Continuous variables are presented as the mean ± standard deviation or median (interquartile range) according to the distribution. Categorical variables are presented as frequencies and percentages. Continuous variables were compared using Student’s *t*-test or Mann–Whitney U-test. Categorical variables were compared using the chi-square or Fisher’s exact test. Event rates for MACE were calculated using the Kaplan–Meier method and the log-rank test was used to compare events across atrial GLS groups based on the optimal cut-off obtained from the receiver operating characteristic curve to predict MACE. Univariate and multivariate Cox proportional hazard regression analyses were performed to clarify variables related to the occurrence of MACE. Although the limited number of events restricted the inclusion of covariables in the multivariate model, multivariate models were constructed, including age, sex, New York Heart Association class, and atrial GLS. The results are reported as hazard ratios with 95% confidence intervals. A p-value of < 0.05 was considered statistically significant. All statistical analyses were performed using the Statistical Package for the Social Sciences software (version 26; IBM Corp., Armonk, NY, USA) and the R statistical package (version 4.1.1; R Foundation for Statistical Computing, Vienna, Austria).

## Results

### Patients’ characteristics and outcomes

We retrospectively analysed 96 patients in the study (61.6 ± 11.3 years old, 37 males). Baseline characteristics of patients with and without MACE, including clinical, laboratory, and echocardiographic data, are shown in Table 1. The prevalence rates of extra-cardiac sarcoidosis involvement and comorbidities were not significantly different between the two groups. Patients with MACE had a higher New York Heart Association class and plasma B-type natriuretic peptide levels than those without MACE. During the follow-up, 85 % of patients received steroids. Some patients had device implantation (pacemaker: 19%, implantable cardioverter defibrillator: 26%, cardiac resynchronization therapy defibrillator: 28%).

**Table 1:**
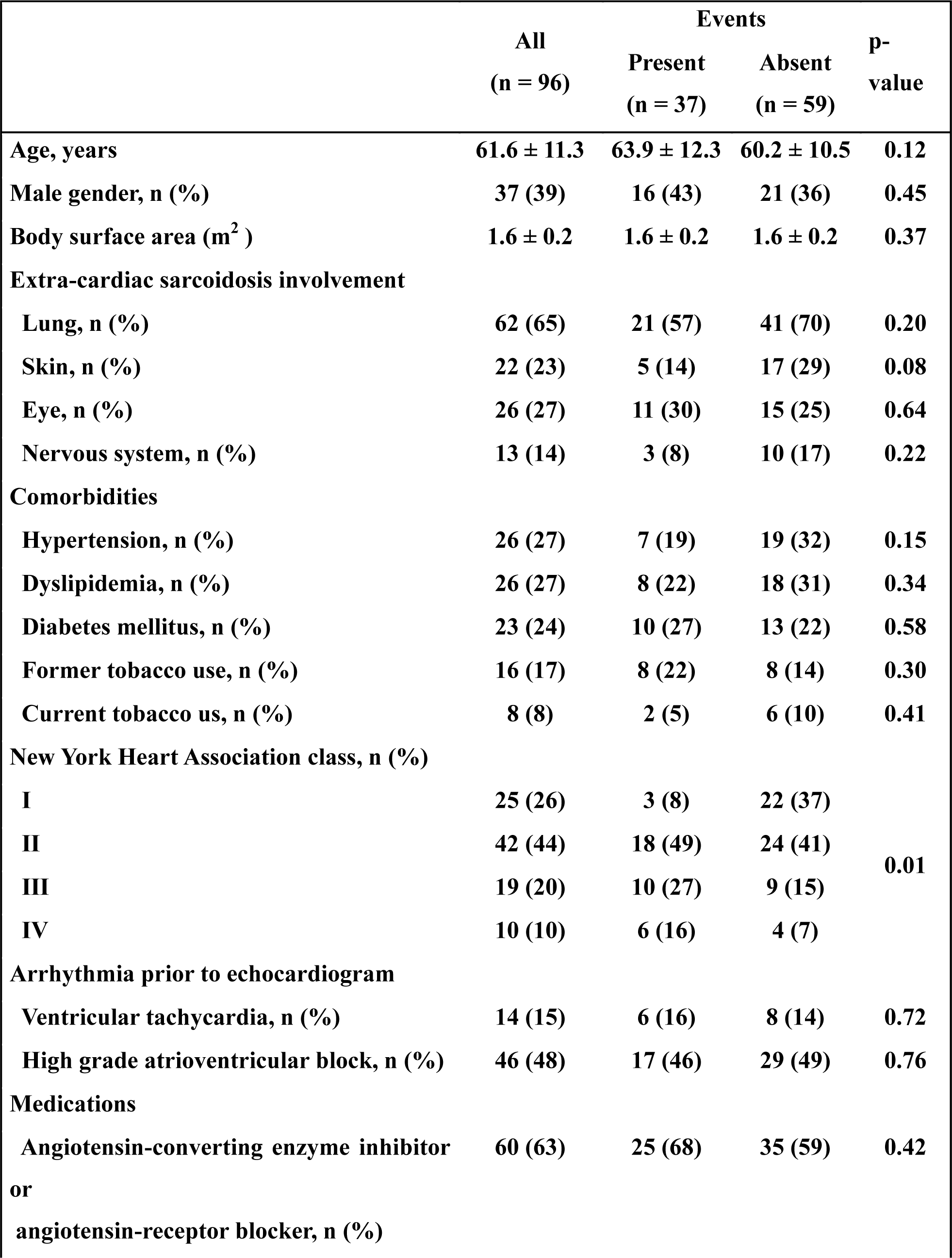
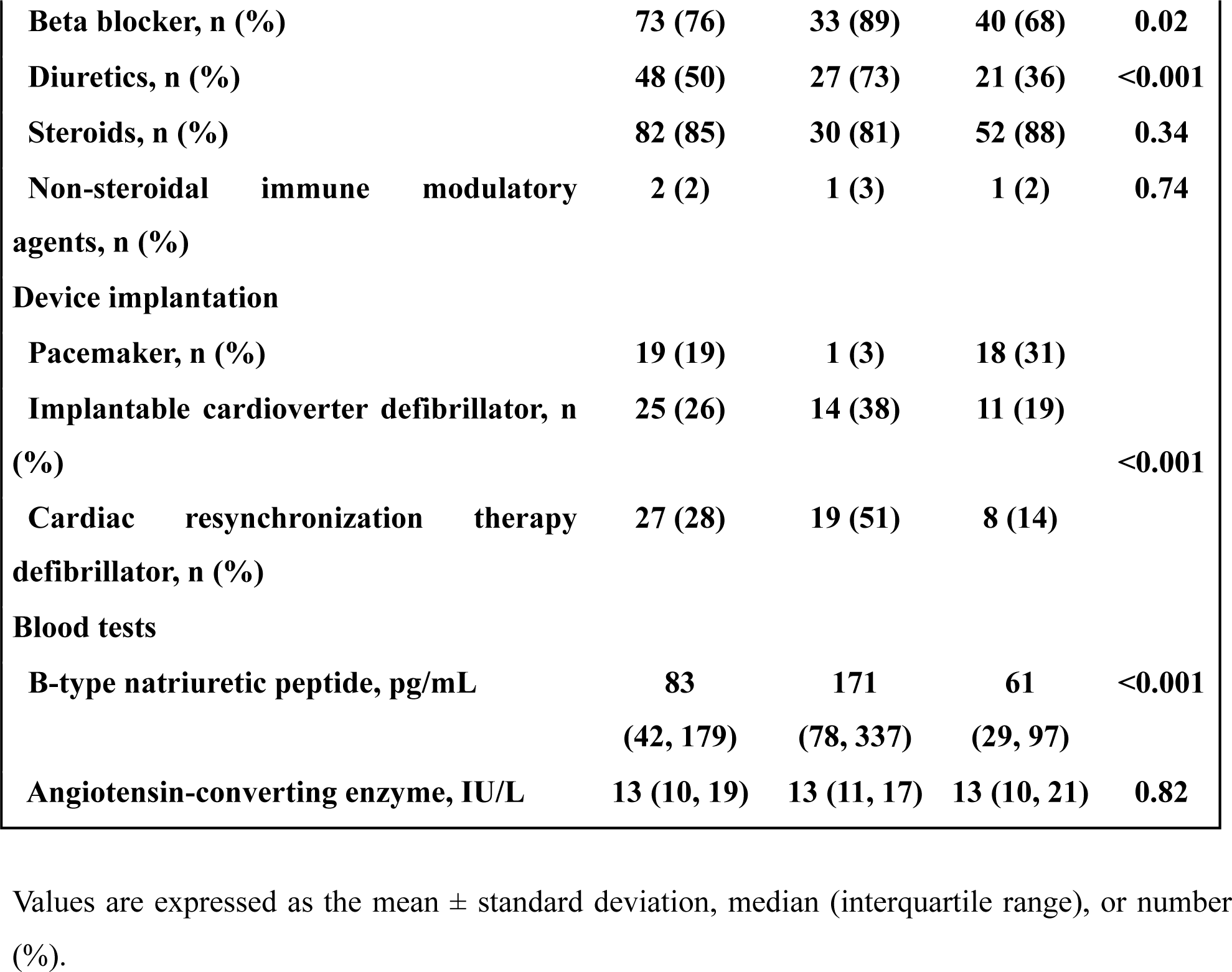
Clinical characteristics of the study population.

During a median follow-up of 6.0 years (interquartile range: 2.9-9.9 years), 37 patients had MACE (7 cases of cardiac death, 23 cases of fetal ventricular arrhythmia events, and 7 cases of hospitalization for heart failure).

### Echocardiographic characteristics

A comparison of echocardiographic characteristics between the two groups is shown in Table 2. Regarding LV function parameters, LV ejection fraction was significantly lower and LV dimension was significantly higher in patients with MACE than in those without MACE. Regarding RV function parameters, patients with MACE had a lower RV fractional area change than those without MACE. Regarding atrial function parameters, LAGLS and RAGLS were significantly lower in patients with MACE than in those without MACE (14 ± 8% vs 22 ± 9%, p < 0.001 and 18 ± 9% vs 25 ± 11%, p < 0.001, respectively). LA volume index and RA volume index were also significantly higher in patients with MACE than in those without MACE.

**Table 2:**
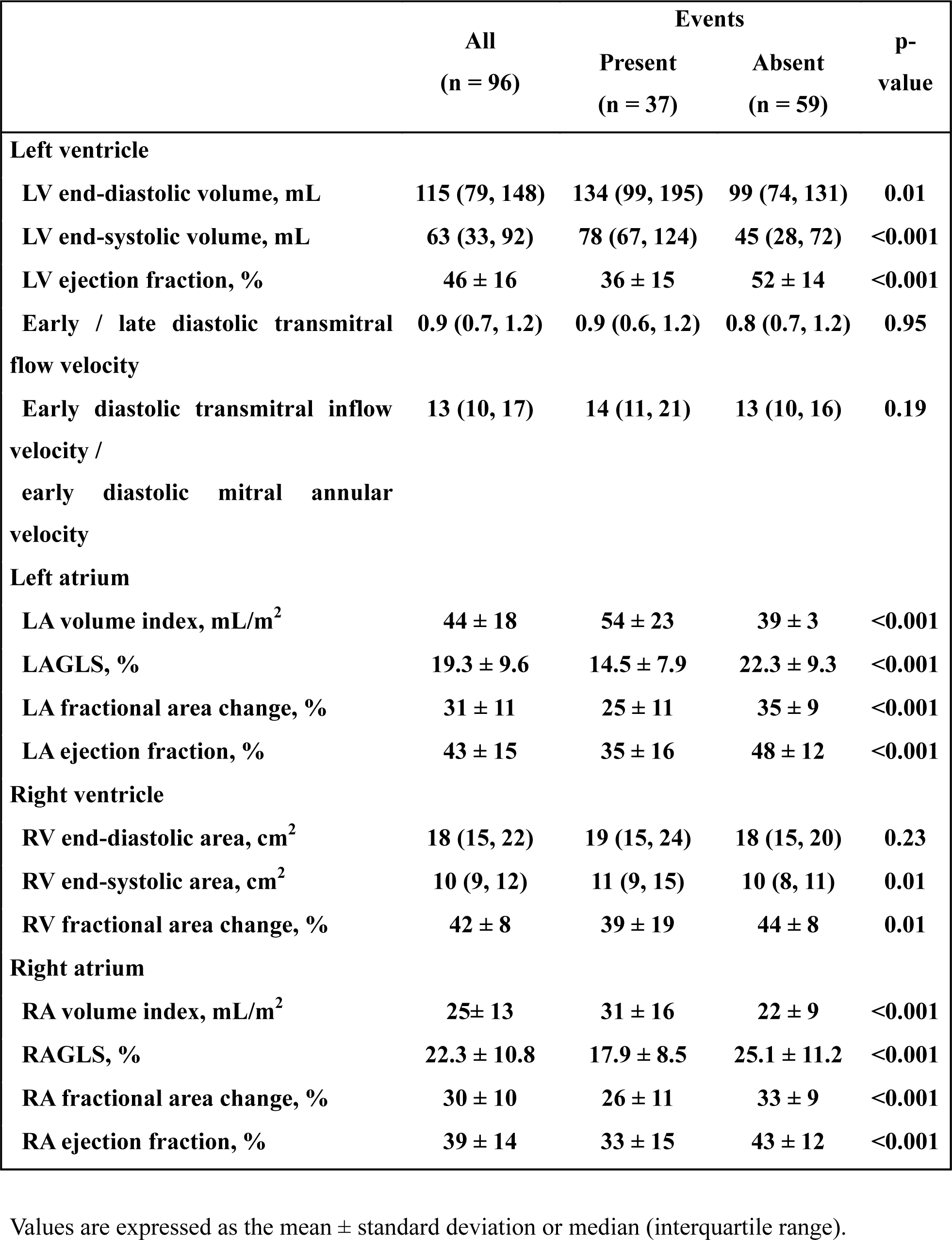
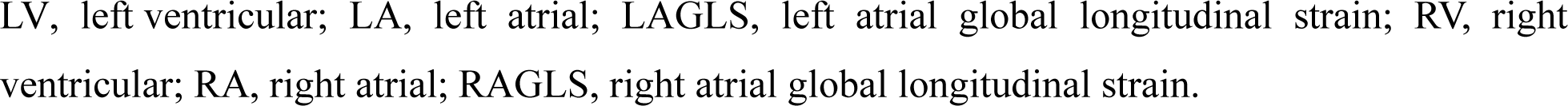
Echocardiographic characteristics.

### Association of atrial function with clinical outcomes

The corresponding optimal cut-off values of LAGLS and RAGLS for discriminating MACE from the receiver operating characteristic curve were 19.6% and 22.3%, respectively. The cumulative incidence of MACE was significantly higher in patients with low LAGLS of ≤ 19.6% than in those with high LAGLS of > 19.6% (log-rank test, p = 0.01, Figure 3A). The cumulative incidence of MACE was significantly higher in patients with low RAGLS of ≤ 22.3% than in those with high RAGLS of > 22.3% (log-rank test, p = 0.01, Figure 3B).

**Figure 3.**
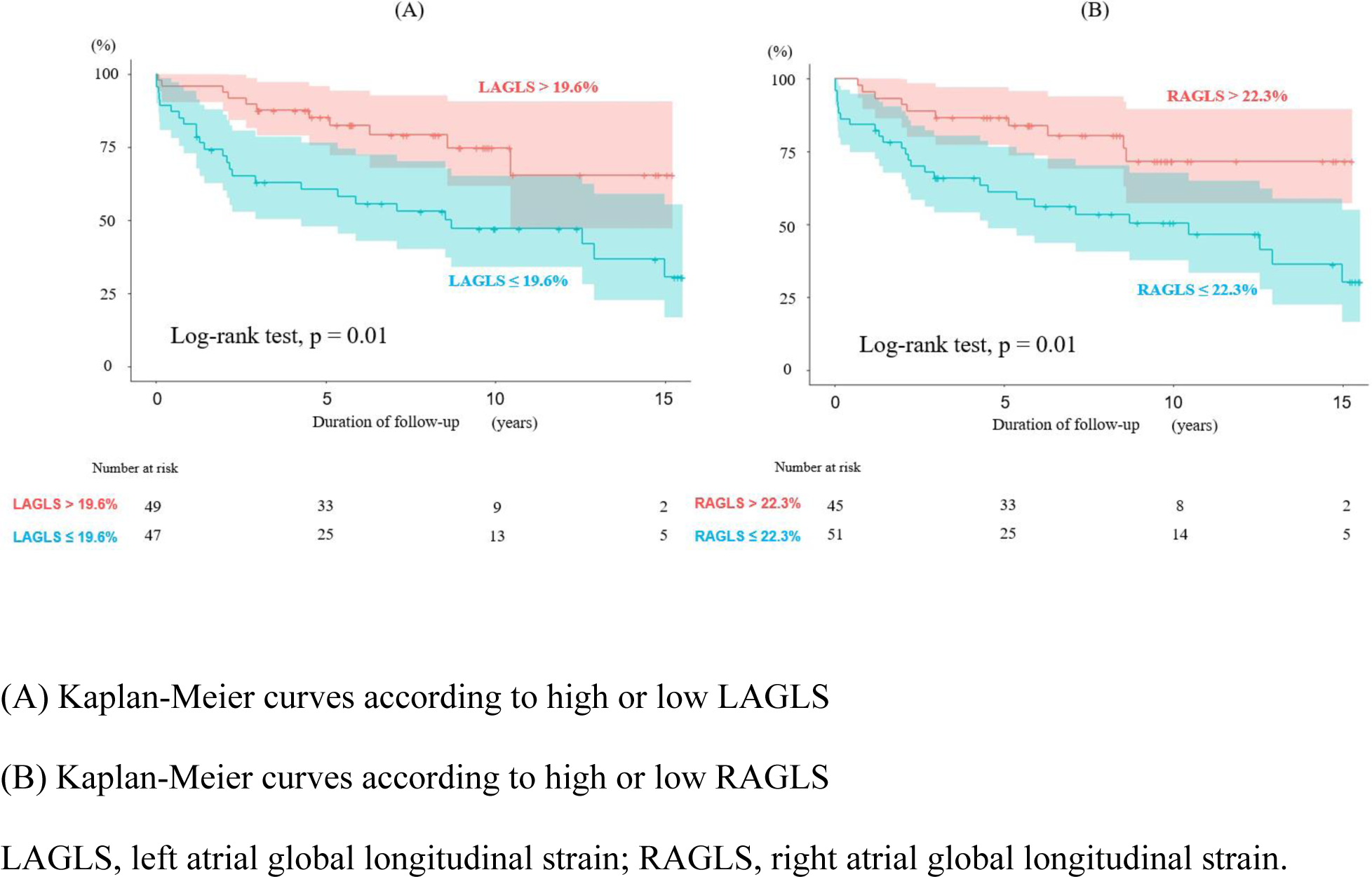
Kaplan-Meier curves of cumulative probability of event-free survival according to atrial function.

Cox proportional hazards regression analyses showed that age ≥ 60 years, New York Heart Association class ≥ Ⅲ, LV ejection fraction ≤ 40%, RV fractional area change ≤ 35%, LAGLS ≤ 19.6%, and RAGLS ≤ 22.3% were significantly associated with MACE in the univariate analysis (Table 3). In multivariate analysis, LAGLS ≤ 19.6% and RAGLS ≤ 22.3% were independently associated with MACE in a model including age ≥ 60 years, sex, and New York Heart Association class ≥ Ⅲ (hazard ratio: 2.19, 95% confidence interval: 1.06–4.55, p = 0.04; hazard ratio: 2.27, 95% confidence interval: 1.07–4.85, p = 0.03, respectively).

**Table 3:** Univariate and multivariate analyses of prediction of the outcome endpoints.

|  | Univariate |  | Multivariate 1 |  | Multivariate 3 |  |
| --- | --- | --- | --- | --- | --- | --- |
|  | Hazard ratio | p-value | Hazard ratio | p-value | Hazard ratio | p-value |
|  | (95% confidence interval) |  | (95% confidence interval) |  | (95% confidence interval) |  |
| Age | 1.86 | 0.08 | 2.02 | 0.06 | 1.81 | 0.11 |
| (≥ 60 years) | (0.93-3.74) |  | (0.98-4.15) |  | (0.87-3.78) |  |
| Male | 1.44 | 0.27 | 1.65 | 0.14 | 1.47 | 0.27 |
|  | (0.75-2.77) |  | (0.85-3.21) |  | (0.75-2.87) |  |
| New York Heart Association class | 2.37 | 0.01 | 2.22 | 0.02 | 2.42 | 0.01 |
| (≥ III) | (1.24-4.56) |  | (1.13-4.38) |  | (1.25-4.69) |  |
| LV ejection fraction | 3.67 | <0.001 |  |  |  |  |
| (≤ 40%) | (1.89-7.1) |  |  |  |  |  |
| RV fractional area change (≤ 35%) | 2.50 | 0.01 |  |  |  |  |
|  | (1.25-5.00) |  |  |  |  |  |
| LAGLS | 2.72 | 0.01 | 2.19 | 0.04 |  |  |
| (≤ 19.6%) | (1.34-5.53) |  | (1.06-4.55) |  |  |  |
| RAGLS | 2.67 | 0.01 |  |  | 2.27 | 0.03 |
| (≤ 22.3%) | (1.30-5.60) |  |  |  | (1.07-4.85) |  |

LV, left ventricular; RV, right ventricular; LAGLS, left atrial global longitudinal strain; RAGLS, right atrial global longitudinal strain.

## Discussion

The present study showed that reduced LAGLS and RAGLS were associated with adverse outcomes, including cardiac death, fetal ventricular arrhythmia events, and hospitalization for heart failure, in patients with CS. To the best of our knowledge, this is the first study to show the importance of atrial function in relation to the prognosis in patients with CS.

### Ventricular function and CS

Cardiac involvement with sarcoidosis is associated with an increased risk of mortality due to congestive heart failure and ventricular tachyarrhythmias ^2–4^ ^14^. The most prognostic factor in CS has been recognized as LV dysfunction ^15^ ^16^. A retrospective nationwide study reported that LV dysfunction predicted poor long-term outcomes, such as cardiac death and transplantation, in 110 patients with CS ^15^. A retrospective multicenter registry of 512 patients with CS reported that LV ejection fraction independently predicted adverse outcomes defined as all-cause death, hospitalization for heart failure, or documented fetal ventricular arrhythmia events ^16^. In addition to LV function, RV function has recently been the focus of attention because of advances in imaging modalities. In one study of 290 patients with CS, RV dysfunction measured by cardiac magnetic resonance was independently associated with all-cause death ^7^. In one study of 899 patients with CS, RV ejection fraction and RV late gadolinium-enhancement assessed by cardiac magnetic resonance were associated with a composite outcome of all-cause death, cardiovascular events, or sudden cardiac death ^8^.

### Atrial function and CS

CS affects various parts of the myocardium. In autopsy studies, Roberts et al. reported that sarcoidosis lesions were found not only in the LV free wall, the ventricular septum, and the RV wall, but also in the RA and the LA ^17^. Previous studies reported atrial involvement in approximately 20% of patients dying from or with CS ^18^ ^19^. These findings indicate that sarcoidosis develops in the atrium as well as in the ventricle. Recently, the relationship between cardiac involvement and supraventricular arrhythmias has been mentioned. Viles-Gonzalez et al. reported that 32% of patients with CS experienced supraventricular arrhythmias, including atrial fibrillation, atrial tachycardias, atrial flutter, and other types of supraventricular tachycardias ^20^. Supraventricular arrhythmias are thought to be caused by atrial inflammation and scarring and atrial overload secondary to ventricular dysfunction or pulmonary lesions ^21^. Habibi et al. reported that atrial inflammation detected by fluorine-18 fluorodeoxyglucose positron emission tomography and atrial remodeling assessed by cardiac magnetic resonance were associated with the occurrence of atrial fibrillation ^22^. Niemelä et al. also reported that atrial uptake of fluorine-18 fluorodeoxyglucose on positron emission tomography was a predictor of subsequent atrial fibrillation ^23^. Atrial remodeling due to inflammation and scarring induces atrial fibrillation, leading to the development of impairment of atrial function. Atrial dysfunction plays an important role in cardiac outcomes ^24^. However, the prognostic significance of atrial function has not been evaluated in patients with CS.

### GLS using 2D-STE

Two-dimensional-STE is an innovative method to identify subclinical myocardial damage and detect systolic dysfunction in regional myocardial deformation at an early stage of the disease ^25^. Recently, in patients with heart failure, the evaluation of atrial function has become useful in predicting the prognosis ^26^ ^27^. Carluccio et al. reported that LAGLS of ≤ 12.9% was associated with adverse outcomes, such as all-cause death and hospitalization for heart failure, in patients with heart failure with reduced ejection fraction ^26^. Fread et al. reported that LAGLS of ≤ 31.2% was associated with worse outcomes in patients with heart failure with preserved ejection fraction ^27^. Regarding GLS in CS, a few studies reported the usefulness of 2D-STE for assessing LV function to predict the prognosis in sarcoidosis ^28–30^. Cristina et al. reported that reduced LVGLS and RVGLS were correlated with adverse cardiac events ^30^. However, no studies have investigated the effect of atrial GLS on clinical outcomes in patients with CS.

The present study showed that LAGLS and RAGLS were associated with cardiac death, fatal ventricular arrhythmia events, and hospitalization for heart failure during the long-term follow-up. Our findings suggest that atrial function has prognostic value in patients with CS. The evaluation of atrial function could be useful for identifying risk stratification in patients with CS. In this study, the cut-off values of LAGLS and RAGLS to show the accuracy to predict events were ≤ 19.6% and ≤ 22.3%, respectively. These cut-off values were not clearly defined and could vary by the disease. Further studies are required to investigate the disease-specific optimal cut-off of LAGLS and RAGLS for identifying clinical outcomes.

### Study limitations

The present study had some limitations. First, this study was conducted at a single center with a limited number of events. Large studies are required to confirm our findings. However, this study included a relatively large number of patients with CS despite it being a rare disease. Second, this study did not assess the interaction of clinical outcomes with therapeutic management. Finally, risk stratification of CS by cardiac magnetic resonance has been previously mentioned, but this imaging modality was not assessed in this study.

## Conclusions

Atrial dysfunction represented by LAGLS and RAGLS using 2D-STE is associated with adverse outcomes in patients with CS. Our findings suggest that atrial function has prognostic value in patients with CS. The assessment of atrial function could be effective for identifying patients at a high risk and for considering appropriate therapeutic strategies.

## Data Availability

None declared.

## Acknowledgments

All authors appreciate medical technologists for their efforts in measuring atrial global longitudinal strain.

